# ADHD is more closely linked to neurodevelopmental than externalizing and internalizing disorders: A genetically informed multivariate Swedish population study

**DOI:** 10.1101/2020.02.26.20028175

**Authors:** Ebba Du Rietz, Erik Pettersson, Isabell Brikell, Laura Ghirardi, Qi Chen, Catharina Hartman, Paul Lichtenstein, Henrik Larsson, Ralf Kuja-Halkola

**Affiliations:** Department of Medical Epidemiology and Biostatistics, Karolinska Institutet, Stockholm, Sweden; The National Centre for Register-based Research, Department of Economics and Business Economics, Business and Social Science. Aarhus University, Aarhus, Denmark; Department of Psychiatry, University of Groningen, University Medical Center Groningen, Groningen, The Netherlands; School of Medical Sciences, Örebro University, Örebro, Sweden

## Abstract

**Background:** While ADHD is currently classified as a neurodevelopmental disorder in the latest diagnostic manuals, the disorder shows phenotypic and genetic associations of similar magnitudes across neurodevelopmental, externalizing and internalizing disorders. This study aimed to investigate if ADHD is etiologically more closely related to neurodevelopmental than externalizing or internalizing disorder clusters after accounting for a general psychopathology factor.

**Methods:** Full- and maternal half-sibling pairs (N=774,416), born between 1980 and 1995, were identified from the Swedish Medical Birth and Multi-Generation Registers, and ICD-diagnoses were obtained using the Swedish National Patient Register. A higher-order confirmatory factor analytic model was performed to examine associations between ADHD and a general psychopathology factor as well as a neurodevelopmental, externalizing, and internalizing subfactor. Quantitative genetic modelling was performed to estimate the extent to which genetic, shared and non-shared environmental effects influenced the associations with ADHD.

**Results:** ADHD was significantly and strongly associated with all three neurodevelopmental, externalizing and internalizing factors (r=0.67-0.75). However, after controlling for a general psychopathology factor, only the association between ADHD and the neurodevelopmental-specific factor remained moderately strong (r=0.43, 95%Confidence Interval [CI]=0.42-0.45) and was almost entirely influenced by genetic effects. In contrast, the association between ADHD and the externalizing-specific factor was smaller (r=0.25, 95%CI=0.24-0.27), and largely influenced by non-shared environmental effects. There remained no internalizing-specific factor after accounting for a general factor.

**Conclusions:** ADHD comorbidity is largely explained by genetically influenced general psychopathology, but the strong link between ADHD and other neurodevelopmental disorders is also substantially driven by unique genetic influences.

## Introduction

Attention-deficit/hyperactivity disorder (ADHD) is a common psychiatric disorder, with a prevalence of 3.4-7.1% in childhood and adolescence (1,2) and 2.8-3.4% in adulthood (3-5). Genetic factors contribute strongly to ADHD liability; twin and family studies have estimated the heritability of ADHD at 70-80% (6-10) and measured common genetic variants explain 22% of phenotypic variance in ADHD (11).

In the former versions of the diagnostic manuals Diagnostic and Statistical Manual of Mental Disorders (DSM-IV-TR) and International Statistical Classification of Diseases and Related Health Problems (ICD-10) (12,13), ADHD was classified as part of *disruptive behavior disorders*, often referred as ‘externalizing’ disorders (14,15). In the recently updated versions of DSM-5 and ICD-11 (16,17), ADHD is now classified as a *neurodevelopmental disorder*, together with Autism Spectrum Disorder (ASD), disorders of intellectual development, learning disorders, and motor disorders. This change is supported by the high comorbidity and similar characteristics between ADHD and neurodevelopmental disorders, in terms of implicated neurocognitive impairments and early disorder-onset (18-20).

Research, however, suggests substantial genetic overlap between ADHD and not only neurodevelopmental disorders, but also externalizing and internalizing disorders (11,21,22). A recent meta-analysis of twin studies showed that pooled genetic correlations between ADHD and neurodevelopmental, externalizing and internalizing traits were of similar magnitude; estimates ranged from 0.49-0.56, with overlapping confidence intervals (CIs) (21). These quantitative genetic findings have been paralleled by recent molecular genetic research, further suggesting similar magnitudes of genetic associations between ADHD and neurodevelopmental, externalizing and internalizing disorders (11,22,23). One important finding to note in molecular genetic studies is however that the strongest genetic correlation with ADHD has often been reported for depression (11,22). This might, however, reflect statistical power, as the genome-wide association study on depression is by far the largest among psychiatric disorders. Quantitative genetic studies using multivariate approaches may be useful to accurately compare the strengths of associations between psychiatric disorders.

The substantial degree of genetic overlap between psychiatric disorders demonstrates that genetic variants that contribute to risk for developing psychiatric disorders have highly pleiotropic effects, i.e., influencing multiple disorders. There has been an increased research interest into a latent general psychopathology (P) factor that explains phenotypic and genetic variation (10-57%) across psychiatric conditions, including ADHD (24-32). However, it remains unclear whether genetic effects for ADHD are shared with other neurodevelopmental disorders, as well as externalizing and internalizing disorders, after accounting for general psychopathology. Some evidence for genetic specificity comes from a population-based study that investigated associations between polygenic risk score (PRS) for ADHD and psychiatric childhood symptoms after controlling for a general P factor (27); the specific association between ADHD PRS and hyperactivity-impulsivity symptoms was significant, but not with other neurodevelopmental, externalizing or internalizing symptoms (27). This finding is largely in line with another study using ADHD PRS in children (33). These prior studies, however, used PRSs of limited power to capture genetic variation associated with disease specificity (27,33,34), and have mainly focused on psychiatric symptoms obtained in childhood. Further research, using statistically powerful methods and clinical assessments beyond childhood, are therefore needed to understand if ADHD is etiologically closer to certain psychiatric domains after accounting for general psychopathology.

The aim of this study is to use a large-scale multivariate sibling design of children and young adults, followed until 18-33 years, to estimate the general versus specific (after controlling for general effects) psychopathological influences that explain the associations between ADHD and comorbid disorder clusters (neurodevelopmental, externalizing and internalizing), and to what extent these are genetic or environmental in origin. Despite uncertainties in the current research literature, we hypothesize that ADHD will be most etiologically closely linked to the neurodevelopmental cluster after accounting for a general P factor, in light of the similar disorder characteristics (18-20), and which would support the structure of the recently revised diagnostic manuals.

## Methods and Materials

### Sample

Our source population consisted of all individuals born in Sweden 1980-1995 identified from the Medical Birth Register (1,688,807 individuals). We excluded individuals who were born with congenital malformations (80,912 excluded), died (7,412 excluded) or emigrated (57,389 excluded) before the age of 15. Using Swedish personal identification numbers, we linked several nation-wide registers. The National Patient Register (NPR) includes psychiatric inpatient admissions in Sweden since 1973 and outpatient diagnoses since 2001, classified according to the ICD version eight (1969-1986), nine (1987–1996) or ten (1997–present). Life-time diagnoses were treated as binary variables (presence/absence). Each participant could receive more than one diagnosis during the study period.

Using the Multi-Generation Register we identified all full and maternal half-siblings, excluding adopted children (11,266 excluded). Among individuals without half-siblings, we selected one random full sibling pair per family who were not twins. In families with half-siblings, we randomly selected one pair of maternal half-siblings. In total, we included 774,416 individuals from 341,066 full- and 46,142 maternal half-sibling pairs. The study had ethical approval from the Regional Ethical Review Board in Stockholm, Sweden (Dnr 2013/862–31/5).

### Measures

We extracted main and secondary ICD (versions 9 and 10) diagnoses from inpatient or outpatient services from the NPR. We focused on neurodevelopmental, externalizing and internalizing disorders that are polygenic in nature (e.g., Rett’s syndrome was excluded; Table S1 for included disorders and ICD-codes).

### Statistical analysis

#### Phenotypic analyses

We grouped the disorders based on the structure in DSM-5 and ICD-11, and on clustering structures from previous literature (14,35-37). The neurodevelopmental subfactor included ASD, developmental and learning disorders, intellectual disability and motor disorders. The externalizing subfactor included oppositional defiant disorder [ODD], conduct disorder [CD], antisocial personality disorder, alcohol misuse, and drug misuse. CD, ODD and antisocial personality disorders were grouped as one disorder category due to low prevalence rates and the moderately high conversion rate of ODD/CD into adult antisocial personality disorder (38,39). The internalizing subfactor included depression, general anxiety, phobic disorders, reactions to severe stress and adjustment disorders (e.g., post-traumatic stress disorder), and obsessive-compulsive disorder (OCD).

We fitted a higher-order confirmatory factor analytic model on individual-level phenotypic data across full- and maternal half-sibling pairs; henceforth referred to as the **general factor model** (Figure 1). This model was suitable for our research question, as it enables partitioning the relative contributions of general and cluster-specific factors from the overall correlations between ADHD and disorder clusters. Psychiatric disorders were set to load onto one of the three latent subfactors, henceforth referred to as the neurodevelopmental, externalizing, and internalizing subfactors. We modelled a general P factor, which loaded onto each of the three subfactors. Correlations between the subfactors were fixed to zero so that they only correlated through the P factor. Each subfactor had an additional loading from a specific factor (i.e., residual variance not explained by the general factor), henceforth referred to as the neurodevelopmental-, externalizing-, and internalizing-specific factors. Correlations between each specific factor and the general factor were fixed to zero. We then correlated ADHD with the general and the three specific factors.

**Figure 1.**
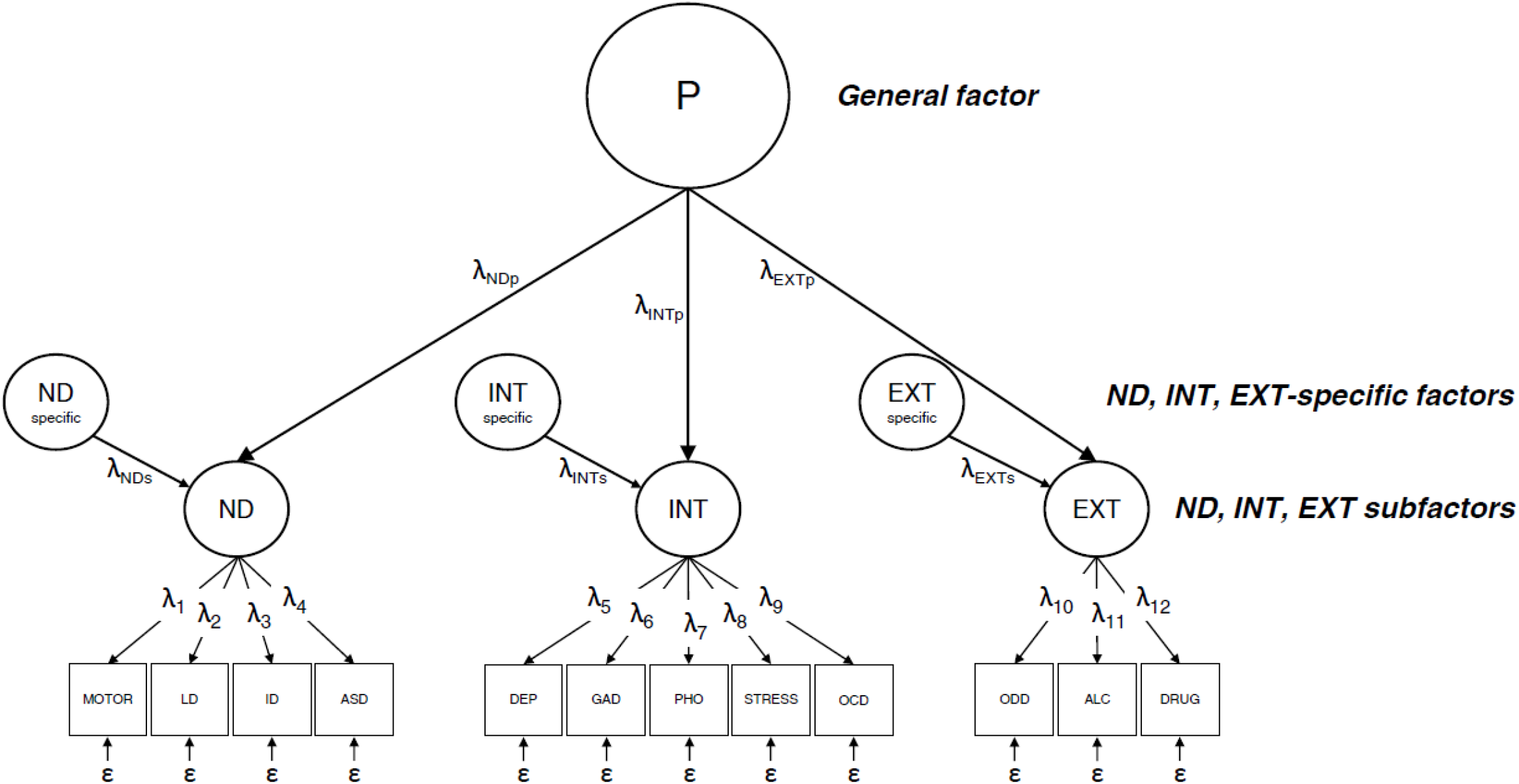
General factor model. Note: P; General psychopathology factor, ND; Neurodevelopmental subfactor, INT; Internalizing subfactor, EXT; Externalizing subfactor, ND specific; Neurodevelopmental-specific factor, INT-specific; Internalizing-specific factor, EXT-specific; Externalizing-specific factor, ADHD; Attention-deficit/hyperactivity disorder, Motor; Motor disorders, LD; Learning and developmental disorders, ID; Intellectual disability, ASD; Autism spectrum disorder, DEP; Depression, GAD; Generalized anxiety disorders, PHO; Stress; Reactions to severe stress and adjustment disorders, OCD; Obsessive-compulsive disorder, ODD; Oppositional defiant, conduct and antisocial personality disorders, ALC; Alcohol misuse, DRUG; Drug misuse.

#### Quantitative genetic analyses

We used a multivariate sibling design to establish the genetic and environmental etiology of the general factor and the three subfactors (neurodevelopmental, externalizing, internalizing) by specifying additive genetic (A), shared environmental (C), and non-shared environmental (E) latent variables as sources of variance and covariance between factors (25,40). A-variables were estimated by fixing them to correlate between siblings at their expected average sharing of co-segregating genes (0.5 for full siblings, 0.25 for half-siblings). C-variables (i.e., non-genetic components that make sibling pairs similar) were estimated by fixing them to correlate at unity across full and half-siblings. Thus, we assumed the shared environment variables to be equally shared across full and maternal half-siblings, as previous literature has shown strong support for this assumption in Swedish registers (25). E-variables were estimated by fixing them to correlate at zero across all siblings, thus measuring non-genetic components making siblings within a pair dissimilar.

All analyses included sex and birth year (as categories, see Table 1) as covariates to adjust for different follow-up lengths and cohort effects. Analyses were performed in R (version 3.4.1) using the ‘corrplot’ (version 0.84) and ‘OpenMx’ packages (version 2.15.5) (41,42). Because half-siblings tend to display a higher rate of disorders than full siblings (43,44), prevalence rates were allowed to vary across full and half-siblings. We used weighted least squares for model fitting, and χ^2^-tests to compare nested models. We evaluated if the models provided a good fit using the root mean square error of approximation (RMSEA; comparison to the maximum possible fit to the data) and the comparative fit index (CFI; comparison to a model where correlations between observed variables are assumed to be zero) (45). Data analysis was performed between May 3, 2019, and January 14, 2020.

**Table 1.**
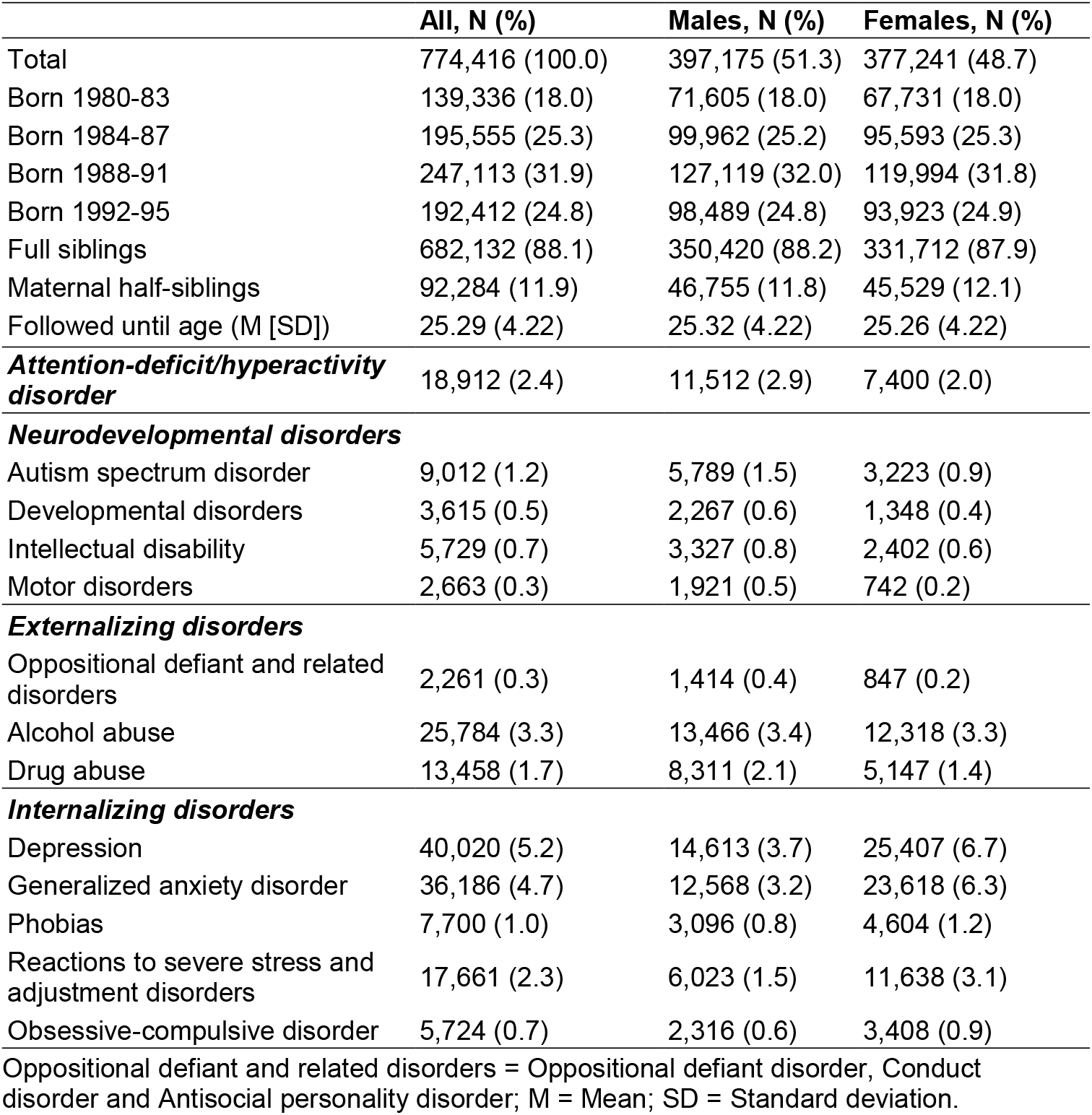
Descriptive statistics in study sample: age, sex and prevalence rates of psychiatric disorders

|  | **All, N (%)** | **Males, N (%)** | **Females, N (%)** |
| --- | --- | --- | --- |
| Total | 774,416 (100.0) | 397,175 (51.3) | 377,241 (48.7) |
| Born 1980-83 | 139,336 (18.0) | 71,605 (18.0) | 67,731 (18.0) |
| Born 1984-87 | 195,555 (25.3) | 99,962 (25.2) | 95,593 (25.3) |
| Born 1988-91 | 247,113 (31.9) | 127,119 (32.0) | 119,994 (31.8) |
| Born 1992-95 | 192,412 (24.8) | 98,489 (24.8) | 93,923 (24.9) |
| Full siblings | 682,132 (88.1) | 350,420 (88.2) | 331,712 (87.9) |
| Maternal half-siblings | 92,284 (11.9) | 46,755 (11.8) | 45,529 (12.1) |
| Followed until age (M [SD]) | 25.29 (4.22) | 25.32 (4.22) | 25.26 (4.22) |
| ***Attention-deficit/hyperactivity disorder*** | 18,912 (2.4) | 11,512 (2.9) | 7,400 (2.0) |
| ***Neurodevelopmental disorders*** |  |  |  |
| Autism spectrum disorder | 9,012 (1.2) | 5,789 (1.5) | 3,223 (0.9) |
| Developmental disorders | 3,615 (0.5) | 2,267 (0.6) | 1,348 (0.4) |
| Intellectual disability | 5,729 (0.7) | 3,327 (0.8) | 2,402 (0.6) |
| Motor disorders | 2,663 (0.3) | 1,921 (0.5) | 742 (0.2) |
| ***Externalizing disorders*** |  |  |  |
| Oppositional defiant and related disorders | 2,261 (0.3) | 1,414 (0.4) | 847 (0.2) |
| Alcohol abuse | 25,784 (3.3) | 13,466 (3.4) | 12,318 (3.3) |
| Drug abuse | 13,458 (1.7) | 8,311 (2.1) | 5,147 (1.4) |
| ***Internalizing disorders*** |  |  |  |
| Depression | 40,020 (5.2) | 14,613 (3.7) | 25,407 (6.7) |
| Generalized anxiety disorder | 36,186 (4.7) | 12,568 (3.2) | 23,618 (6.3) |
| Phobias | 7,700 (1.0) | 3,096 (0.8) | 4,604 (1.2) |
| Reactions to severe stress and adjustment disorders | 17,661 (2.3) | 6,023 (1.5) | 11,638 (3.1) |
| Obsessive-compulsive disorder | 5,724 (0.7) | 2,316 (0.6) | 3,408 (0.9) |

Oppositional defiant and related disorders = Oppositional defiant disorder, Conduct disorder and Antisocial personality disorder; M = Mean; SD = Standard deviation.

#### Secondary analyses

To allow comparability with prior findings in the literature, we ran additional analyses using methodological approaches that have been more commonly used in research. First, we conducted univariate ACE modeling of the psychiatric disorders to allow comparability to estimates from twin studies. Second, we conducted a correlated factors model, where disorders loaded onto one of the three neurodevelopmental, externalizing and internalizing subfactors (allowing subfactors to correlate and without a general factor). We additionally ran a bifactor model, which derives the P factor from the correlation matrix between the psychiatric disorders rather than higher-order subfactors. With this bifactor model, we aimed to investigate if the patterns of within-individual and between-sibling correlations of ADHD with the cluster-specific factors remained when the correlation between disorders in different clusters were not forced to correlate through P via the cluster subfactors. To examine the robustness of our findings across different sibling selection criteria, we re-ran the general factor quantitative genetic analyses on sibling pairs born closest together rather than random pairs in a family.

## Results

In the sample, 51.3% were males, and the average follow-up length was until 25.3 years of age (Table 1 for information on sex, birth year and prevalence of psychiatric disorders). Figure 2 displays the within-individual phenotypic (tetrachoric) correlations between psychiatric disorders (Table S2 for 95%CIs). The overall pattern of correlations showed that psychiatric disorders most strongly correlated with other disorders within their respective, pre-specified neurodevelopmental, externalizing or internalizing clusters, while the correlations between ADHD and other disorders were difficult to characterize.

**Figure 2.**
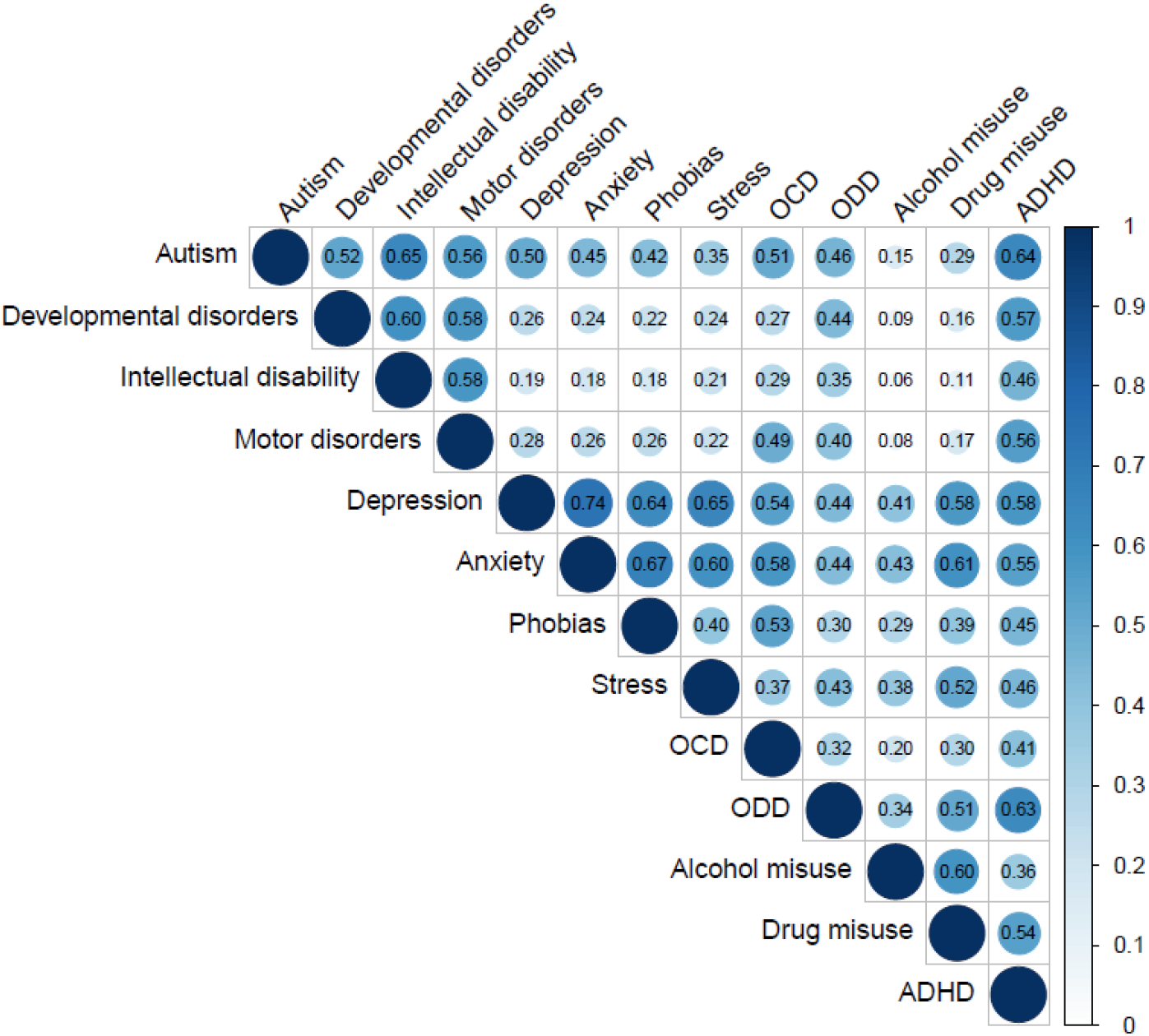
Observed within-individual phenotypic correlations between disorders. Note: OCD; Obsessive-compulsive disorder, ODD; Oppositional defiant disorder, conduct disorder and antisocial personality disorder, ADHD; Attention-deficit hyperactivity disorder.

### General factor model

#### Phenotypic analyses

The general factor model fit the data well (RMSEA=0.006, CFI=0.974). The neurodevelopmental, externalizing and internalizing subfactors all loaded moderately to strongly on the P factor (factor loadings 0.60, 0.77, & 0.99, respectively) (Table S3a). ADHD was significantly and strongly correlated with the P factor (r=0.67). The total correlations between ADHD and the neurodevelopmental, externalizing and internalizing subfactors (sum of contributions through P and cluster-specific factors) were 0.75, 0.67 and 0.67, respectively. Because the internalizing subfactor loaded almost perfectly on the P factor, we could not estimate valid standard errors. We therefore fixed the internalizing subfactor to have a loading of 1 on the P factor, which did not lead to a worse model fit (Table S3b), and did not affect the subsequent estimates (Table S3-8). In this model, the magnitude of loadings of the neurodevelopmental and externalizing subfactors on the P factor, and their correlations with ADHD, remained unchanged (Figure 3, Tables S3c & S6 for 95%CIs).

**Figure 3.**
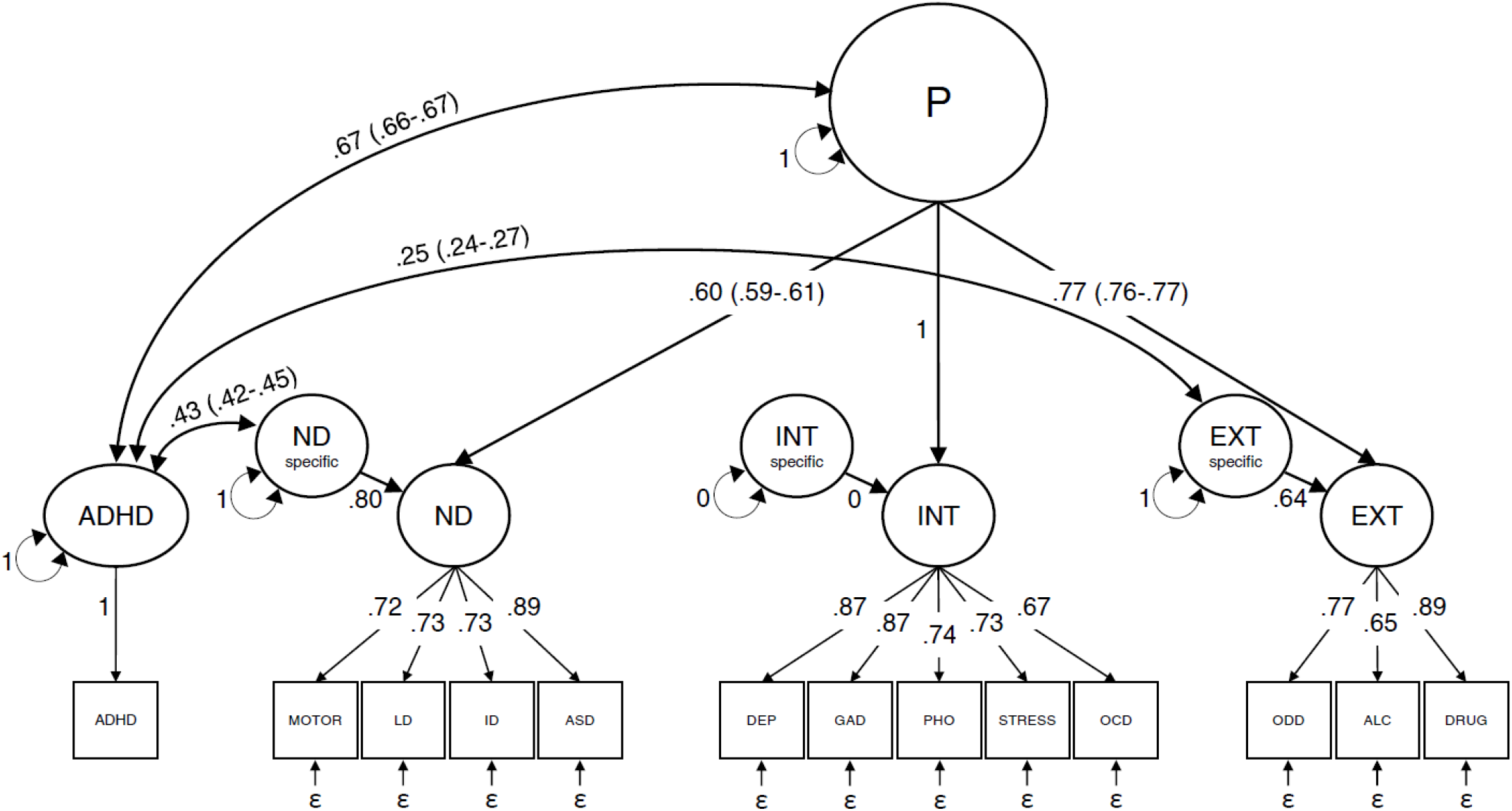
General factor solution: factor loadings and phenotypic correlations between ADHD and the general psychopathology (P) factor, neurodevelopmental-specific and externalizing-specific subfactors. Note: P; General psychopathology factor, ND; Neurodevelopmental subfactor, INT; Internalizing subfactor, EXT; Externalizing subfactor, ND specific; Neurodevelopmental-specific factor, INT-specific; Internalizing-specific factor, EXT-specific; Externalizing-specific factor, ADHD; Attention-deficit/hyperactivity disorder, Motor; Motor disorders, LD; Learning and developmental disorders, ID; Intellectual disability, ASD; Autism spectrum disorder, DEP; Depression, GAD; Generalized anxiety disorders, PHO; Stress; Reactions to severe stress and adjustment disorders, OCD; Obsessive-compulsive disorder, ODD; Oppositional defiant, conduct and antisocial personality disorders, ALC; Alcohol misuse, DRUG; Drug misuse.

After accounting for the general P factor, ADHD showed a significant and moderately strong phenotypic correlation with the neurodevelopmental-specific factor (r=0.43, 95%CI=0.42-0.45), and a significantly smaller correlation with the externalizing-specific factor (r=0.25, 95%CI=0.24-0.27). Given that variance in the internalizing factor was entirely subsumed by its covariance with the P factor (loading fixed at 1), a correlation was not estimable with the internalizing-specific factor (Figure 3 & Table S6).

#### Quantitative genetic analyses

Observed correlations among psychiatric disorders and latent factors between full- and maternal half-sibling pairs are shown in Tables S7 and S9.

The model fitting results estimated the heritability of the general psychopathology factor at 0.49, with the contribution of shared environment at 0.07 and non-shared environment at 0.44. For the neurodevelopmental-specific and externalizing-specific factors, the estimated heritabilities were 0.89 and 0.80, the shared environment contributions 0.09 and 0.06, and the non-shared environment contributions 0.03 and 0.14, respectively. Estimates are plotted in Figure 4 and corresponding 95%CIs are reported in Table S10 (see Table S11 for ACE correlations).

**Figure 4.**
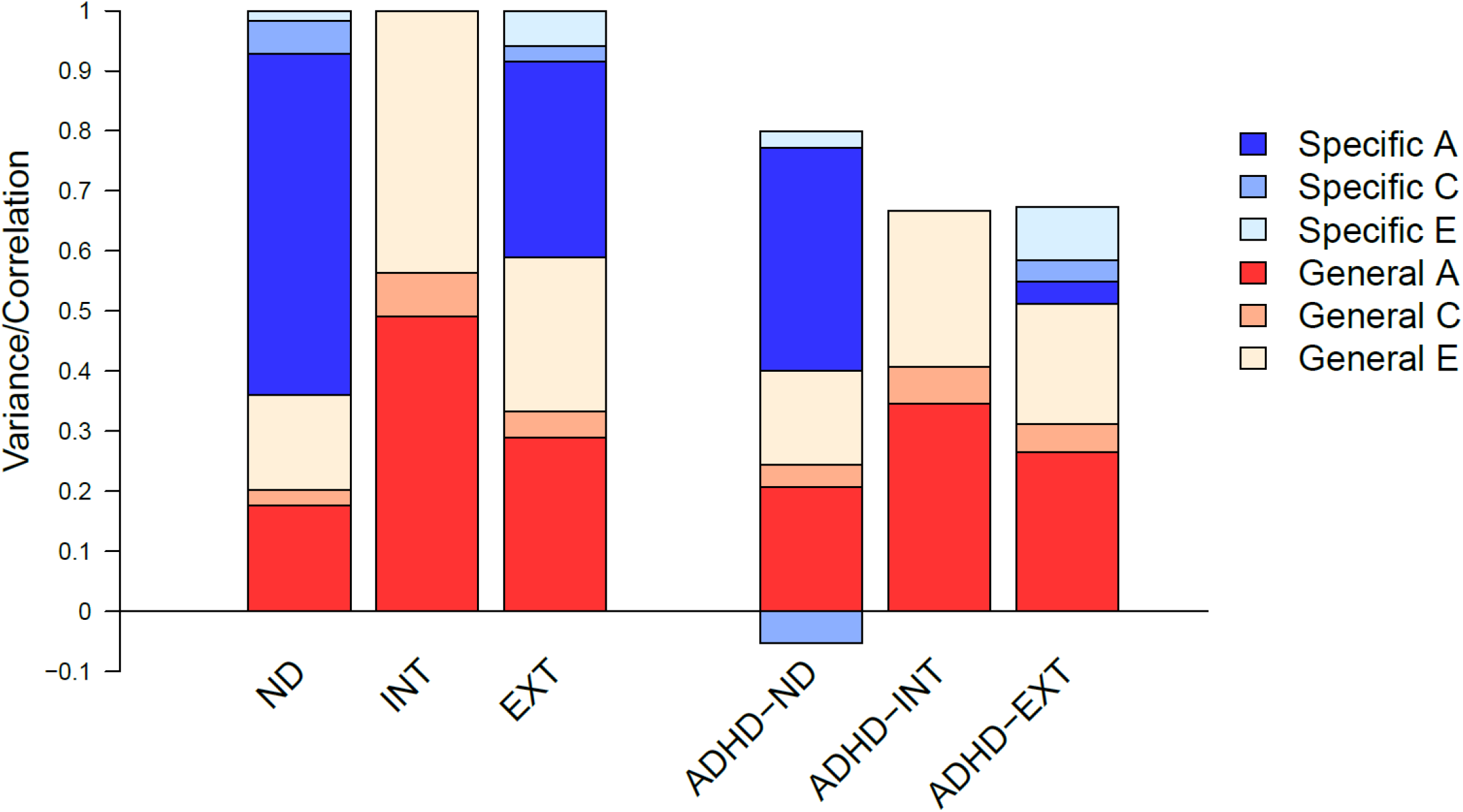
General factor solution: proportion of variance in the subfactors, and in their phenotypic correlations with ADHD, explained by specific and general additive genetic (A), specific and general shared (C) and specific and general non-shared (E) environmental effects. Note: ADHD; Attention-deficit/hyperactivity disorder, ND; Neurodevelopmental, INT; Internalizing, EXT; Externalizing, See Figure S14 for estimates and 95% Confidence Intervals.

For the phenotypic correlation between ADHD and the P factor (r=0.67), genetics contributed with 0.35 (52% of total correlation), shared environment contributed with 0.06 (9%) and non-shared environment contributed with 0.26 (39%) (Table S10 for estimates and 95%CIs). For the phenotypic correlation between ADHD and the neurodevelopmental-specific factor (r=0.43), genetics contributed with 0.46 (107%; greater than 100% due to negative contribution to correlation from shared environment), shared environment contributed with -0.07 (-15%; negative contribution to correlation), and non-shared environment contributed with 0.04 (8%).

For the phenotypic correlation between ADHD and the externalizing-specific factor (r=0.25), genetics contributed with 0.06 (23%), shared environment contributed with 0.06 (22%), and non-shared environment contributed with 0.14 (55%) (Figure 4, Table S10).

### Secondary analyses

See Table S12 for univariate ACE model estimates for single disorders, which are overall in line with prior estimates from twin studies (7,10,46-48). The correlated factors model (RMSEA=0.006, CFI=0.974) revealed that the three neurodevelopmental, externalizing and internalizing subfactors were moderately-to-strongly intercorrelated (r=0.46-0.77), and were all significantly and strongly correlated with ADHD (r=0.67-0.75). The magnitude of correlations, along with the heritabilities and co-heritabilities of the subfactors and ADHD, were overall in line with previous research (Figure S1) (11,21,24,27,35,49,50). The bifactor model (RMSEA=0.004, CFI=0.982) revealed very similar within-individual and between-sibling correlations of ADHD with the general and specific factors compared to the higher-order model (Figure S2 & Table S13).

When we re-ran analyses on siblings born closest together, the estimates and model fit remained very similar to when we used random sibling pairs in a family (Figure S3). The negative contribution of the shared environment on the correlation between ADHD and the neurodevelopmental-specific factor decreased slightly in the analyses of siblings born closest together.

## Discussion

In this large-scale, register-based sibling study we showed for the first time using clinical diagnoses that ADHD is more phenotypically and genetically linked to neurodevelopmental disorders than to externalizing and internalizing disorders, after accounting for a general P factor.

The phenotypic correlations between ADHD and the neurodevelopmental, externalizing and internalizing subfactors were all strong, and were moderately-to-strongly influenced by genetic variation, in line with previous research (21,30,49,50). After accounting for the P factor, the correlation between ADHD and the neurodevelopmental-specific factor remained moderately strong, and was largely genetic in origin, suggesting substantial unique sharing of biological mechanisms between disorders. In contrast, the correlation between ADHD and the externalizing-specific factor was much smaller and was largely explained by non-shared environmental effects. The observation that there remained an association between ADHD and the externalizing-specific factor after accounting for familial (genetic and shared-environmental) effects, is in line with a causal framework of ADHD on externalizing disorders. ADHD, which often has an early onset, might subsequently lead to later-onset substance misuse or antisocial personality disorder. Additional research, using longitudinal causal modelling approaches, may further unravel the effect of ADHD on subsequent externalizing disorders. Lastly, the correlation between ADHD and the internalizing subfactor was almost entirely explained by the P factor. This finding suggests that the comorbidity of ADHD and internalizing disorders is largely due to pleiotropic genetic effects and non-shared environmental influences that have general effects on psychopathology.

### Research and clinical implications

While past research, and results from our correlated factors model, suggest similar magnitudes of genetic overlap between ADHD and different psychiatric disorders, we found that after accounting for general psychopathology, the strongest genetic sharing was with neurodevelopmental disorders. These findings demonstrate that isolating the P factor may allow for identifying specific mechanisms (e.g., biological pathways) and risk factors, uniquely related to only a subset of disorders or syndromes, and may have large potential for future etiological research. For example, the Hierarchical Taxonomy of Psychopathology framework hypothesizes that a higher-order approach to phenotypes may increase the precision of molecular genetic findings, by differentiating between genetic liability for broad psychopathology and dimension-specific genetic risk factors.^51^

Our findings on the close unique genetic link between ADHD and the neurodevelopmental subfactor are in line with the current diagnostic classification of ADHD as a neurodevelopmental disorder, despite that we used ICD-codes from the former classification system. We acknowledge that there is debate regarding to what extent genetic structure of diseases should be used to inform diagnostic nosology (52-54). However, a common view in research is that genetic data, and mechanisms learned from them, can, at least in part, aid in the formulations/revisions of the clinical syndromes and diagnostic classification (54).

## Limitations

This large-scale study has several strengths, including the use of comprehensive and clinical assessments of psychiatric disorders across childhood and young adulthood, and the use of a representative population cohort. However, findings should be interpreted in light of some limitations.

Individuals with multiple psychiatric diagnoses or with siblings who have diagnoses may be more likely to get in contact with the mental health system, which can lead to an overestimation of associations among disorders and sibling pairs. Nevertheless, we note that our results of a general factor of psychopathology are largely consistent with previous survey studies, suggesting that results are not entirely due to these biases (27,30).

During the follow-up period of individuals in our cohort, there have been changes in diagnostic practices for several psychiatric disorders (e.g., the rate of ADHD diagnoses increased fivefold from 2004 to 2015) (55), and the register coverage has improved. Further, there were variations in follow-up length between individuals in our study. To account for these issues, we adjusted for the association between birth year and the expected prevalence for each disorder. However, if the change in diagnostic practices and register coverage has in turn changed the phenotypes and etiologies of the diagnoses, or have had differential effects on the disorders, we may have remaining bias in our results. Replication using other data sources, or similar sources with longer follow-up, would strengthen the inferences from the current study. For the ACE models, we assumed that full- and maternal half-siblings share their common (family) environment to the same degree. We acknowledge that this is a simplification, but sensitivity analyses in a similar study showed strong support for this assumption (25). Further, when we re-ran analyses with siblings born closest together, i.e. siblings who are closer in age and may therefore share more common environment, our overall results remained unchanged.

Lastly, our finding that the internalizing subfactor loaded nearly perfectly on the P factor has been reported in previous research (30), and studies have often reported that internalizing disorders have the, or among the, highest loadings on the general factor (24,26,31,34). However, it is possible that this high loading may have partly resulted from the pre-defined confirmatory approach to factor definitions. Furthermore, the chosen disorders in a study dictates what the P factor captures, thus, the resulting structure will be partly specific to the current study design. The bifactor model produced very similar within-individual and between-sibling correlations between ADHD and the neurodevelopmental, externalizing and internalizing subfactors compared to the higher-order model. This supports the robustness of our results, and suggests that the almost perfect loading of the internalizing subfactor on P is not a major source of bias.

## Conclusion

ADHD comorbidity is largely explained by partly genetically influenced general psychopathology, but the strong link between ADHD and other neurodevelopmental disorders is also driven by specific genetic influences. These findings support the classification of ADHD as a neurodevelopmental disorder in the recently revised diagnostic manuals, and provide insights into the structure of the etiologic underpinnings of psychopathology.

## Data Availability

Data cannot be shared publicly due to the Swedish Secrecy Act. Data from the Medical Birth Register, the Multi-Generation Register, and the National Patient Register were used for this study and made available by ethical approval. Researchers may apply for access through the Swedish Research Ethics Boards (www.etikprovningsmyndigheten.se) and from the primary data owners Statistics Sweden (www.scb.se), and the National Board of Health and Welfare (socialstyrelsen.se), in accordance with Swedish law.

## Acknowledgements

Henrik Larsson acknowledges financial support from the Swedish Research Council (2018-02599) and the Swedish Brain Foundation (FO2018-0273). Ebba Du Rietz was supported by grant 2019-01172 from the Swedish Research Council for Health, Working Life, and Welfare. Erik Pettersson was supported by grant 2017-01358 from the Swedish Research Council.

